# Anxiety and media exposure during COVID-19 outbreak in Kuwait

**DOI:** 10.1101/2020.08.24.20180745

**Authors:** Lulua Alasousi, Sara al Hammouri, Sara al Al-abdulhadi

## Abstract

**Background:** Rising fear and panic among public during COVID19 pandemic increase concern regarding anxiety cases in Kuwait. Media capture our attention during this period looking for daily virus update lead to more fear. Our purpose of this study to examine the relationship between anxiety and media exposure among Kuwaiti during COVID19 outbreak

**Method:** cross sectional study among Kuwaiti citizen between age23-55yrs old was conducted from April,21,2020 to May,15,2020 using online survey. Total of 1230 participants involve in the current study after exclusion criteria removed. Beside demographic data and media exposure anxiety was assessed using generalized anxiety disorder scale GAD-7, multivariable regression was used to identify the correlation between anxiety and media exposure

**Result:** the result show that there is positive correlation between media exposure and anxiety during COVID19 outbreak in Kuwait (p<.001), furthermore it revealed that there is significant relationship between the frequency of exposure and anxiety(<.001)

**Conclusion:** from this study we can understand that during COVID19 pandemic exposure to media can cause anxiety therefore measures should be taken by the governments to fight misinformation and physician should pay more attention to mental health disease during this period.

## Background

The 2020 coronavirus outbreak has changed lives around the world. It impacted even the minute details of our daily activities making it a health crisis worth attention. A growing body of literature suggests that disasters can have health consequences for the victims involved, such as posttraumatic stress disorder, depression, anxiety, or substance abuse(1,2) Increased self-reports of nonspecific psychological distress and medically unexplained physical symptoms (e.g., fatigue, head-ache, difficulty concentrating, joint/muscle pain) have been noted following disasters as well. Studies conducted during and after epidemics such as SARS of 2003 and Ebola of 2014, observed that there was a widespread fear overactive behavior among the general public(3,4)

The infection caused by the novel coronavirus (COVID-19) was first declared in December 2019 in Wuhan, Hubei, Province China. Since then, more cases were identified in a growing number of international locations. The World Health Organization has declared the current outbreak of COVID19 as pandemic on March,11,2020 (5)which lead to the activation of emergency state around the world. The first known case of COVID19 was reported in Kuwait on Feb,24,2020(6) By May 22, 2020, 19,564 confirmed COVID19 cases were announced (6)After SARS and MERS, COVID-19 has been the third pandemic caused by the coronavirus(7) that has led to worldwide panic due to it is a rapid increase in cases and deaths.

In response to this pandemic, strict preventative measures were implemented such as shelter in place and the closure of shops, schools, and government institutions. Studies show that social isolation, long and strict curfew measure, economic fall and media lead to increase psychological distress during pandemic(8)

Perhaps one of the major drivers of physiological distress is the media. Unsurprisingly, Covid19 outbreak attracted copious media attention. Both traditional (T.v and newspaper) and nontraditional media (twitter and WhatsApp) were involved in covering the pandemic around the globe. During a health crisis, the public depends on the press to convey accurate and up to date information and shape public perception around health issue, to make informed decisions regarding health-protective behaviors(9-11). Public health offices and governments using these channels to communicate effectively with public. Decision science has revealed that people tend to form accurate perceptions of risk when facts are known and communicated to the public effectively via the media (12,13)

Research shows that media has a major impact on mental health and physiological behavior. Studies linked media violence to antisocial behavior (14)and ideal body deception to body image disturbance and eating disorders(15) Media framing can shape how audiences feel or think about an issue(16,17)

It has a tone of voice, supported by the use of fearful or reassuring words and metaphors (18) A content analysis shows that the evolution of information from press release to news is marked by significant changes in media frames (19)In pandemics, catastrophe events with potentially large impacts but low probability of occurrence of which individuals have little direct cognitive experience, media frames may have an even more pronounced effect on their audiences (20,21)

Media also played a role in the emotional response to the H1N1 outbreak. Media “hype” and uncertainty of information have caused panic at the beginning of the pandemic. Media hype is defined as Extravagant or intensive publicity created by or by means of the mass media, especially out of proportion to the person or thing being publicized. However, a few months into the pandemic when vaccine became ready for distribution, the severity of the disease was deemed to be mild and a lack of compliance with recommended preventive behaviors prevailed(22)

In this day and age, there is a 24-hour news cycle readily available in the palm of our hands. It allows people to view stories from all over the world and stories to be repeated and exaggerated. Media coverage of the pandemic is broadcasted from international locations transmitting either reassurance or distress. For example media coverage of Italian army carrying dead bodies due to the high number of deaths on that day went viral all over the world(23). Studies show that traumatic images on the media will evoke negative emotional responses to the viewer(24) Also, repeated media exposure to information about an infectious disease particularly can exacerbate stress responses, worry, and impair functioning(25)

Social media is a chief part of the news coverage available to the public. Social media platforms act as a primary bridge between individuals and the news sources, aggregating traditional and social media into one convenience feed(26). News is shared through the eyes of the individual and their emotions of anxiety or reassurance can be translated to the viewing public. While some accounts may be official and verified, most social media accounts are not subject to verification and may broadcast questionable information that is subject to the individuals credibility.

In Kuwait, social media is a powerful communication platform. In 2019, 3.9 million users (93% of the total population) were documented around the country(27) Many users resorted to the social media to express their feelings during this pandemic, many of whom are doctors and health officials. Other than social media, since the start of the pandemic our official media was very active in providing the latest news regarding the new cases and death through our daily press conference(6)

The aim of this study was to analyze the media role in the mental health response, especially anxiety, to the pandemic. As family physicians, we are faced with many vague symptoms that are diagnosed as anxiety during this health crisis. By understanding the media role, we can form a complete view of the drivers of anxiety for our patients and address how to respond to the news in our management plans.

## Method

The study is a cross-sectional observational study conducted from April 21 2020 to May 15 2020 during the covidl9 outbreak in Kuwait (done during partial curfew). The study included Kuwaiti residence from the age of 23-55 year and excluded the following:

1. citizens who are under home or institutional quarantine
2. COVID19 patient
3. doctors, nurse or pharmacist who work in the ministry of health in Kuwait
4. cancer, psychiatric disease, diabetes mellitus, and hypertension patient
5. non-Kuwaiti citizens

Ethical committees approved this study in the ministry of health Kuwait (1409/2020). An online semi-structured questionnaire was developed with a consent form attached to it. It was designed to be answered only once. The survey included the following:

1) demographic data:

the information of basic demography was collected such as age, gender, marital status, educational level, employment, and governorates

2) information about media exposure:

media exposure was measured by asking: over the last week, how many time you follow covidl9 news ? Response options were: “less” once a day,” sometimes” half of the day and “frequently” all-day

3) anxiety assessment:

GAD-7 score was adopted (28)comprising seven items to test. Translation into Arabic was done in progressive steps to ensure that the essential meaning was preserved. The original questionnaire in English were translated into Arabic by bilingual Arabic speaking family physician. Another Arabic speaking family physician independently performed translation back into English, which was again retranslated into Arabic. Participants were asked how often they were bothered by each symptom during the last two weeks. Response options were “not at all,” several days,” more than half of the days” and “nearly every day,” scored as 0,1,2 and 3 respectively. A score of more than 4 was consider as anxiety (5-9 mild anxiety, 10-14 moderate anxiety, and 15-21 severe anxiety) according to the GAD-7 system.

### reliability testing

Table 1 represents the results of the reliability analysis a particular item with the sum of the rest of the items. While the values in the column labeled as “Cronbach’s alpha if item dropped” are the values of overall alpha if that item is not included in the calculation.

**Table (1):** Instruments reliability analysis using Cronbach’s Alpha

|  | mean | SD | item-rest correlation | If item dropped |
| --- | --- | --- | --- | --- |
| | | | | Cronbach's $\alpha$ |
| employment status | 2.203 | 1.830 | -0.057 | 0.899 |
| feeling nervous, anxious or on the edge | 1.046 | 0.934 | 0.556 | 0.732 |
| not able to stop or control worrying | 0.777 | 0.935 | 0.672 | 0.714 |
| worrying too much about different things | 1.129 | 0.958 | 0.681 | 0.711 |
| unable to relax | 0.958 | 0.999 | 0.643 | 0.716 |
| so restless that it is hard to stay still | 0.526 | 0.839 | 0.681 | 0.717 |
| becoming easily annoyed or irritable | 0.885 | 0.943 | 0.734 | 0.703 |
| feeling afraid as if something awful might happen | 0.711 | 0.936 | 0.568 | 0.730 |

*All Anxiety instruments Cronbach's $\alpha = 0.76$*

The results show that the calculated value of Cronbach’s alpha for all anxiety instruments were 0.769, indicating good reliability, which shows that the questionnaire is reliable and has good internal consistency. Further, it can be seen that the value of “Cronbach’s *α* if item dropped” correspond to the variable “employment status” was higher than calculated value of Cronbach alpha for all anxiety instruments, therefore this item should be removed from the questionnaire before further analysis. Deletion of this item from the questionnaire will increase the value of Cronbach’s alpha from 0.77 to 0.90.

Moreover, corresponding to rest of the items, the value of “Cronbach’s *α* if item dropped” is less than the calculated value of Cronbach’s alpha for all anxiety instruments and also the value of item-rest correlation for these items were more than 0.20, therefore these items should be kept in the questionnaire for further analysis.

#### Pilot

The questionnaire was pretested on sample of 100 persons on April 14 2020 to uncover any difficulties in understanding the meaning of the questions and to estimate the amount of time it would take to complete. Accordingly, some items were modified to improve the participant comprehension. This way, the answers could be standardized and were designed and appended to GAD-7. The results of the pilot showed that 97% of the participants who follow the news have anxiety.

#### Data collection

A snowball sampling technique was used. In total, 3428 participants took part in the survey. After removing the participants who did not meet the criteria, 1230 participants from all the governorates in Kuwait were included in the study. An Arabic and English version was sent via a hyperlink through WhatsApp to contacts of the investigators. The participants were encouraged to roll out the survey to as many people as possible thus the link was forwarded to people apart from the first point of contact and so on. Upon receiving the message and clicking the link, the participants are directed to the information about the study and an informed consent letter.

## Results

All the data was statistically evaluated by using SPSS 25.0 (Statistical Package for Social Sciences). Quantitative variables were represented by mean with standard deviation whereas frequencies and percentages were applied for qualitative variables. Reliability of questionnaire was tested by using cronbach’s alpha coefficient. Further, chi-square (χ^2^) test were applied to observe the association of variables between groups. p-value of less than 0.05 or 0.01 was considered to test the statistical significance of the variables. Finally, logistic regression (univariable and multivariable) was applied to measure the significant effects from study independents variables towards the anxiety levels (yes/no) after adjusting for study confounders.

### 2) Demographic data analysis

Of 1231 participants, 76% were female and 25% were male as it shown in Table 2. Among these, most of the respondents had bachelor degree (61%), followed by respondents having diploma (18%), while 1% of the respondents had less than high school. Further, it can be seen that this study has covered six major governorates of Kuwait, Capital (41%), Hawalli (26%), Mubark al kabeer (12%), Ahmadi (10%), Farwanyia (9%) and Jahra (3%). Majority of the respondents in the survey were married (73%). By considering the age factor, it was found that 41% of the respondents were from age group 34-44, 30% were from age group 23-33 and 29% were from the age-group of 45-55. It can also be observed that more than half of the respondents were government employees (61%) while only 2% of the participants were students.

**Table (2):** Demographics of the study *Summary descriptives table*

|  | [ALL]<br>N=1231 |
| --- | --- |
| Gender: |  |
| Male | 301 (24.5%) |
| Female | 930 (75.5%) |
| Educational level: |  |
| less than high school | 15 (1.22%) |
| high school degree or equivalent | 66 (5.36%) |
| Diploma | 226 (18.4%) |
| bachelor degree | 756 (61.4%) |
| graduate degree | 168 (13.6%) |
| governorates |  |
| Capital | 500 (40.7%) |
| Hawalli | 318 (25.9%) |
| Jahra | 39 (3.17%) |
| Farwanyia | 109 (8.87%) |
| Ahmadi | 118 (9.60%) |
| Mubark al kabeer | 145 (11.8%) |
| Martial status: |  |
| Married | 894 (72.6%) |
| Single | 236 (19.2%) |
| Divorced | 89 (7.23%) |
| Widowed | 12 (0.97%) |
| Age: |  |
| 23-33 | 374 (30.4%) |
| 34-44 | 505 (41.0%) |
| 45-55 | 352 (28.6%) |
| Employment status: |  |
| government employees | 747 (60.7%) |
| private employees | 174 (14.1%) |
| private business | 57 (4.63%) |
| Retired | 154 (12.5%) |
| not employed | 74 (6.01%) |
| Students | 25 (2.03%) |
|  | [ALL] |
|  | *N=1231* |

### 3) Anxiety and media relation

In Table 3 Chi-square test used to check the association between news item and anxiety levels(no, yes). There were 659 respondents under the yes category, while 572 were under the no category of anxiety level. The results show a significant association of exposure to news with anxiety levels. It can be seen from the results that 55% of respondents who follow the news have anxiety while 31.9% who do not follow the news also exhibit anxiety. Furthermore, of the respondents who had a raised anxiety level, the highest percentage (53%) admitted they follow the news frequently. On the contrary, of those who did not have anxiety, the majority (41%) responded that they follow the new less often defined as once a day. Therefore it can be concluded that there is a significant effect of the COVID-19 news on the anxiety level of the respondents. High frequency of following the news increased the anxiety among respondents.

**Table (3):** Summary descriptives table by groups of ‘Anxiety Group’

| News Item | Anxiety = no<br>*N=572* | Anxiety = yes<br>*N=659* | p-value |
| --- | --- | --- | --- |
| Do you follow the news of the spread of the new virus,<br>Corona virus?: |  |  | <0.001 |

| News Item | Anxiety = no<br>N=572 | Anxiety = yes<br>N=659 | p-value |
| --- | --- | --- | --- |
| Yes | 508 (88.8%) | 629 (95.4%) |  |
| No | 64 (11.2%) | 30 (4.55%) |  |
| Over the last week, how many time you follow COVID-19 news?: |  |  | <0.001 |
| Less | 234 (41.0%) | 152 (23.1%) |  |
| Sometimes | 132 (23.1%) | 157 (23.9%) |  |
| Frequently | 205 (35.9%) | 349 (53.0%) |  |

### 4) Statistical analyses (logistic regression)

The logistic regression model formed by considering the news items and other demographic variables was found to be statistically insignificant (χ^2^=13.88 p=0.085) Table 4, though the results of variable as well as multivariable shows the significant effect of the news items and age on dependent variable anxiety. It can be seen that the respondents who follow the news of the spread of the corona virus felt anxious compare to others who did not follow the news. Besides this, the results also show that the respondents who frequently follow the news over the last week were more likely to have anxiety compare to other respondents (OR=2.62; 95% CI: 2.01-3.43; p<0.001).

**Table (4):** Multivariate analysis using logistic regression to measure news influance on Anxiety level after confounding for demographics!!

| Dependent = Anxiety Groups<br>(yes/no) | Levels | Anxiety =<br>no | Anxiety =<br>yes | OR. univariable. | OR. multivariable. |
| --- | --- | --- | --- | --- | --- |
| Do you follow the news of the spread of the new virus, Corona virus? | Yes | 508 (88.8) | 629 (95.4) |  |  |
|  | No | 64 (11.2) | 30 (4.6) | 0.38 (0.24-0.59, p<0.001) | 0.55 (0.33-0.90, p=0.018) |
| Over the last week, how many time you follow COVID-19 news? | Less | 234 (41.0) | 152 (23.1) | - | - |
|  | Sometimes | 132 (23.1) | 157 (23.9) | 1.83 (1.35-2.50, p<0.001) | 1.73 (1.25-2.40, p=0.001) |
|  | Frequently | 205 (35.9) | 349 (53.0) | 2.62 (2.01-3.43, p<0.001) | 2.42 (1.81-3.25, p<0.001) |
| Gender | Male | 167 (29.2) | 134 (20.3) | - | - |
|  | Female | 405 (70.8) | 525 (79.7) | 1.62 (1.24-2.10, p<0.001) | 1.61 (1.21-2.14, p=0.001) |
| Educational level | less than high school | 8 (1.4) | 7 (1.1) | - | - |
|  | high school degree or equivalent | 41 (7.2) | 25 (3.8) | 0.70 (0.22-2.21, p=0.531) | 0.85 (0.26-2.84, p=0.788) |
|  | Diploma | 111 (19.4) | 115 (17.5) | 1.18 (0.41-3.48, p=0.752) | 1.16 (0.38-3.57, p=0.796) |
|  | bachelor degree | 339 (59.3) | 417 (63.3) | 1.41 (0.50-4.05, p=0.515) | 1.39 (0.47-4.22, p=0.549) |
|  | graduate degree | 73 (12.8) | 95 (14.4) | 1.49 (0.51-4.42, p=0.463) | 1.54 (0.50-4.85, p=0.446) |
| governorates | Capital | 234 (41.0) | 266 (40.4) | - | - |
|  | Hawalli | 137 (24.0) | 181 (27.5) | 1.16 (0.88-1.54, p=0.298) | 1.16 (0.86-1.56, p=0.329) |
|  | Jahra | 17 (3.0) | 22 (3.3) | 1.14 (0.59-2.22, p=0.699) | 1.24 (0.62-2.49, p=0.547) |
|  | Farwanyaia | 51 (8.9) | 58 (8.8) | 1.00 (0.66-1.52, p=0.998) | 1.14 (0.73-1.78, p=0.558) |
|  | Ahmadi | 61 (10.7) | 57 (8.7) | 0.82 (0.55-1.23, p=0.339) | 0.83 (0.55-1.28, p=0.404) |
|  | Mubark al kabeer | 71 (12.4) | 74 (11.2) | 0.92 (0.63-1.33, p=0.646) | 0.89 (0.60-1.32, p=0.571) |
| Marital status | Married | 416 (72.7) | 478 (72.5) | - | - |
|  | Single | 108 (18.9) | 128 (19.4) | 1.03 (0.77-1.38, p=0.833) | 1.01 (0.73-1.39, p=0.970) |
|  | Divorced | 41 (7.2) | 48 (7.3) | 1.02 (0.66-1.58, p=0.933) | 1.05 (0.66-1.67, p=0.850) |
|  | Widowed | 7 (1.2) | 5 (0.8) | 0.62 (0.18-1.96, p=0.420) | 0.55 (0.16-1.81, p=0.327) |
| Age | 23-33 | 177 (30.9) | 197 (29.9) | - | - |
|  | 34-44 | 212 (37.1) | 293 (44.5) | 1.24 (0.95-1.63, p=0.115) | 1.11 (0.82-1.49, p=0.498) |
|  | 45-55 | 183 (32.0) | 169 (25.6) | 0.83 (0.62-1.11, p=0.209) | 0.73 (0.51-1.06, p=0.098) |
| Employment status | government employees | 334 (58.4) | 413 (62.7) | - | - |
|  | private employees | 85 (14.9) | 89 (13.5) | 0.85 (0.61-1.18, p=0.324) | 0.92 (0.65-1.31, p=0.650) |
|  | private business | 26 (4.5) | 31 (4.7) | 0.96 (0.56-1.67, p=0.895) | 1.19 (0.67-2.12, p=0.559) |
|  | Retired | 81 (14.2) | 73 (11.1) | 0.73 (0.51-1.03, p=0.075) | 0.91 (0.60-1.39, p=0.655) |
|  | not employed | 36 (6.3) | 38 (5.8) | 0.85 (0.53-1.38, p=0.517) | 0.97 (0.58-1.62, p=0.907) |
|  | Students | 10 (1.7) | 15 (2.3) | 1.21 (0.54-2.82, p=0.641) | 1.33 (0.56-3.29, p=0.527) |

Number of participants = 1231, Number in model = 1227, Missing = 4, AIC = 1652.2, C-statistic = 0.653, H&L = Chi-sq(8) 13.88 (p=0.085)

Moreover, the effect of the factors educational level, governorates, marital status, age and employment status was found to be statistically insignificant, as p>0.05 corresponding to these factors. Therefore, it can be concluded that the educational level, county, marital status, age and employment status of respondents did not have any influence on the anxiety level of the respondents.

## Discussion

The results of the study revealed statistically significant association between media exposure and anxiety. It can be seen from the study results that the participants who follow the news of the spread of the corona virus frequently felt anxious compared to those who did not follow the news. These findings are consistent with previous studies that illustrated that media can cause mental health problems during crisis, such as studies done on COVID 19 outbreak(29)(30), 11 september attack and Iraq war(31) and the Ebola epidemic(25)

During the covid19 outbreak, the news was framed in an upsetting tone. Stories focused mainly on severe health related issues and economic downfall. For example, some of the phrases used were “covidl9 virus is a killer virus”, “there is no treatment for the virus”, and “ the government can’t pay the salaries”. Many news outlets used professionals in the field of medicine and economy to portray these stories. Evidence shows that experts with alarming message are even more attractive for the media (32,33)These frames will result in increased fear and panic among public leading to feelings of anxiety. This was also seen in the coverage of avian flu and SARS which was often sensationalist, focusing on worst-case scenarios and full of emotionally charged language(34-36) This led to widespread panic and mental disturbance.

As previously mentioned, many Kuwaiti citizens are plugged into the social media outlets. It plays a key role in the stories that reach them and the anxiety they feel in relation to the news. In a study conducted in Wuhan focusing on social media exposure during COVID-19 exposure and mental health (29)social media exposure was positively associated with high odds of anxiety. It ties this to disinformation and false reports. The social media in was flooded with rumors of conspiracy theories, fake stories and rumors that the government is hiding the real numbers of cases and deaths from the public. Many health personalities took to the social media to present information that had minimal evidence further enforcing the disinformation. The WHO also recognizes misinformation as a source of uncertainty and mental disturbance and is working tirelessly to fight this “infodemic”(37)

Social media also has a rule in driving anxiety through propagating sensitized information.

Many Kuwaiti citizens saw the race for masks, toilet paper and hand sanitizers from around the world. This also led to panic in Kuwait and raiding of local supermarkets. Videos found their way to the social media accounts which in turn raised the anxiety level. Images of people who broke quarantine spread like wildfire, adding to the anxiety of the citizens as they were certain that people were purposely dispersing the virus. Moreover, many citizens expressed their negative feelings, such as fear, worry, nervous, anxiety on social media, which are contagious social network (38)

Another finding in this study revealed that anxiety is seen more in people who were frequently exposed to media defined as all day exposure. This is consistent with another study that said that there is high prevalence of mental health problems which is positively associated with frequent SME during the COVID-19 outbreak (29)Repeated high media exposure results in a cycle of distress and worry about the future which is commonly associated with PTSD and anxiety(39). For example, in the aftermath of the September 11th (9/11) terrorist attacks, individuals who perceived the media as a provider of useful information were more likely to consume 9/11-related media coverage, but this media use was associated with increased distress over time (40)

Evidence shows that anxiety increases the influx of worried patients to the emergency department as well as healthcare centers thus placing a burden on the health care system(41). Therefore, measures should be taken to deal with unnecessary worry which is driven from sensitized media. We recommend that the government fight the misinformation by filtering the rumors or correct them through national platforms, these national platforms can also act as a help line where people can discuss their worries and fears anonymously with a medically trained professional. As family physicians, we need to collaborated with psychiatrists to develop national guidelines for anxiety screening and treatment during a crisis that focuses on media usage and recognizes its effect on mental health.

This study was limited by the paucity of information regarding the impact of media on mental health disease during crisis. In addition, Kuwait has been hit with previous crises, such as the Gulf War and the bombing of Alsadig mosque, which were not explored and baseline data was not available as to the effect of media coverage of these disasters on anxiety and mental health. Another limitation is the lack of enough information from this questionnaire to calculate the prevalence of anxiety which might illuminate the current mental health status.

For future study, we recommend exploring the causal relationship by cohort study design for the 31.9% of responders who did not follow the news but have increased levels of anxiety.

## Data Availability

data had been taken from the survey which i conducted through2 phases pilot and real population sample among kuwaiti citizens.

## author contribution

Mr. Ahmad al Saber was involved in the processing and interpretation of the data

## Aknowlegement

Thanks to Dr.huda aldewsan chairman of primary care department in KIMS,, Dr. lina, Dr. malka serour, Dr. adanan alwegian, Dr.bashyer albabteen and Ms, Maryam alhelal for their support and collaboration in this research

